# Variation in urobiome composition over time in asymptomatic individuals with spinal cord injury and disease using intermittent catheterization

**DOI:** 10.64898/2026.03.06.26347815

**Authors:** Rochelle E. Tractenberg, Suzanne L. Groah, Ethan Newcomb, Mark Khemmani, Cara Joyce, Alan J. Wolfe, Christopher Riegner

## Abstract

Urinary tract infection (UTI) remains the most common infectious complication among individuals with neurogenic lower urinary tract dysfunction (NLUTD) due to spinal cord injury or disease (SCI/D). Despite widespread reliance on microbiological and symptom-based criteria for UTI diagnosis, significant ambiguity persists—especially in distinguishing clinically meaningful change from normal variability in urinary analysis results. This uncertainty contributes to overdiagnosis, inappropriate antibiotic use, and antimicrobial resistance. The present study seeks to operationalize “normal variability” of the urinary microbiome (urobiome) among adults with SCI/D. Using repeated samples collected from asymptomatic individuals over time, we analyzed inter- and intra-individual microbial composition to determine stability and fluctuation under baseline conditions. We observed wide intra- and inter-individual variability, substantial overlap between asymptomatic and pre-symptomatic states, and a consistent predominance of genera conventionally labeled as “uropathogens” even in the absence of symptoms. These findings suggest that assumptions drawn from cross-sectional studies—linking particular taxa or diversity values to health or disease—are not supported within individuals over time, at least in people with NLUTD. This study provides a foundation for distinguishing expected variation from those potentially related to infection, supporting development of precision-based diagnostic thresholds. Results offer critical insight into the ecological dynamics of the urobiome among people with NLUTD who are asymptomatic, establishes a methodological precedent for urobiome-informed clinical decision-making in SCI/D populations, and provides a foundation for distinguishing expected variation from those potentially related to infection, supporting development of precision-based diagnostic thresholds. By identifying personalized baselines and patterns of change, we aim to support research designed to obtain actionable information from the urobiome to enhance the accuracy and stewardship of UTI diagnosis and treatment in this high-risk population.

## Introduction

Accurate diagnosis of urinary tract infection (UTI) among individuals with neurogenic lower urinary tract dysfunction (NLUTD) due to spinal cord injury or disease (SCI/D) is a major clinical and scientific challenge(1). Targeted quantitative PCR (qPCR) pathogen panels, expanded quantitative urine culture (EQUC), and 16S rRNA gene sequencing have provided deeper resolution of microbial composition of what is now known to be a dynamic microbial ecosystem in the urinary tract, the urobiome(2). These more advanced assays offer the potential to detect organisms missed by standard urine culture (SUC) and identify microbial or host-response signatures associated with disease risk(3).

Interest in 16S rRNA gene sequencing to characterize the urobiome and explore its potential diagnostic and prognostic relevance has been acute and increasing(4). This work has fueled enthusiasm for defining “healthy” and “unhealthy” urobiome profiles(5). However, often urobiome studies rely on cross-sectional samples and simplified summaries of community composition(6) that may obscure intra-individual temporal variability and/or may not capture intra-individual dynamics or reflect underlying biological states(2).

Cross-sectional approaches, although practical, impose several conceptual and methodological limitations. They treat the microbiome as a static entity, assuming that between-person differences observed at a single time point adequately represent stable biological features within individuals(7). This assumption is especially tenuous in the urinary tract, where microbial abundance and composition can fluctuate rapidly in response to hydration, catheterization, antimicrobial exposure, and hormonal or immune factors(2)(8).Without first characterizing the range of normal within-person variability, it is not possible to determine whether a given microbial profile represents a meaningful deviation or simply a transient state captured at a single moment. Nevertheless, studies have characterized entire groups by the predominance category most common among their members(9) even when a substantial proportion of cases share the same predominance profile(10). This majority-rule summarization collapses a multidimensional and temporally variable microbial landscape into dichotomous group categories, erasing within-group heterogeneity and between-group overlap alike. The result is an inferential slide from observed group-level prevalence differences to claims about biological normality or pathology—a conclusion that cross-sectional data, which have never established what is normal within any individual, cannot support.

Methodologically, group-level comparisons of relative abundance or diversity indices obscure the within-person variance that defines microbial ecology. Even when statistically significant differences between groups are reported, they may arise from aggregation artifacts rather than true biological divergence(11). Interpretation is further complicated by compositional data properties: because 16S rRNA profiles reflect relative, not absolute, abundances, apparent differences may emerge simply from changes in one taxon’s proportion rather than in the absolute abundance of others(12,13). The use of single samples per individual amplifies these issues, preventing assessment of whether observed “differences” are stable, cyclical, or stochastic fluctuations within an individual’s normal range(14,15). Without longitudinal validation, inferences drawn from group means risk overstating the diagnostic or etiologic relevance of specific taxa or urotypes(16–18).

Interpretive errors also arise when statistical associations are treated as evidence of causal or diagnostic relationships. For example, defining “dysbiosis” as deviation from the composition most frequently observed among asymptomatic participants presumes that relative frequency equates to biological optimality, yet longitudinal evidence from other body sites demonstrates that “healthy” microbial configurations are often individually unique and dynamically maintained(14,19). In the urinary tract, limited longitudinal data suggest similar individuality and instability(20). Labeling urotypes as “healthy” or “pathogenic” on the basis of cross-sectional prevalence imposes a static, taxonomic interpretation on a dynamic ecological system. Ecological theory instead defines system health in terms of stability, resilience, and functional redundancy, rather than taxonomic diversity(21) or predominance(22,23). As recently as 2022, differences between cross-sectional 16S results from groups of individuals with and without urological diagnoses were incorrectly characterized as “changes”(7). This exemplifies the error of applying a dynamic label to a static observation. Such interpretive shortcuts propagate an illusion of categorical microbial states that do not reflect individual trajectories over time(24).

To address these conceptual, methodological, and interpretive weaknesses, longitudinal studies are required to quantify normal intra-individual variability and test whether group-level inferences are borne out within individuals. A longitudinal design allows examination of temporal stability, transition probabilities, and the persistence of community structure across repeated samples. In the context of SCI/D, where the underlying neurogenic bladder dysfunction, bladder management method, antibiotic exposure, and host physiology interact to shape microbial dynamics, understanding this variability is essential for distinguishing pathological perturbations from normal fluctuation. Without such evidence, the clinical translation of urobiome findings remains speculative.

The present study establishes a baseline for normal variation and provides an empirical test of assumptions underlying cross-sectional urobiome analyses. We analyzed 16S rRNA profiles from each of 94 individuals with NLUTD secondary to SCI/D, with samples collected at least two weeks apart, all obtained during asymptomatic periods. This design enables direct evaluation of variability in genus-level 16S rRNA findings within individuals and the stability of previously identified urotypes over time. By characterizing intra-individual variability and comparing it with cross-sectional patterns reported in the literature, this study quantifies the extent to which prior group-based inferences about “healthy” or “unhealthy” urobiomes align with longitudinal reality.

## Methods

### Participant Recruitment

This is a prospective cohort study of people ages 18 and older with SCI/D (including SCI, multiple sclerosis, and spina bifida) who use intermittent catheterization for bladder management. IRB approval was obtained from our institution (#1124) and informed consent was obtained from all participants prior to inclusion in the study.

Our exclusion criteria included: 1) use of oral or intravesical prophylactic antibiotics; 2) psychologic or psychiatric conditions influencing the ability to follow instructions; 3) use of oral or IV antibiotics within the past 2 weeks; 4) sexual activity within the previous 72 hours; 5) menstruating at the time of collection; and 6) participation in another study that could confound results.

We designed the study to obtain two urine samples, separated by at least two weeks, from eligible individuals who were asymptomatic at the time of sampling. We defined “asymptomatic” as the absence of any symptoms on the Urinary Symptom Questionnaire for Neuropathic Bladder-Intermittent Catheter version (described below) at the time of the urine sample and for 72 hours afterward. If participants developed any symptom on the USQNB-IC within 72 hours of the sample, a new sample was scheduled. Participation ended with two samples were obtained with no symptoms developing within 72 hours (or the study ended).

### Symptom Assessment

We used the Urinary Symptom Questionnaire for Neuropathic Bladder-Intermittent Catheter version (USQNB-IC(25)), a validated tool designed for use by patients, researchers, and clinicians, to assess symptoms in this population. Participants completed the USQNB-IC at least 4 times: at the time of each urine collection (reporting on symptoms over the prior 3 days), and once on each of the three days after each urine collection to ensure that urine samples were “truly asymptomatic” and not “asymptomatic but about to become symptomatic.” Endorsement of any USQNB-IC symptom within the 3 days prior to the sample collection resulted in rescheduling the sample collection for a later time. If the participant endorsed any of the USQNB-IC symptoms during the 72 hours after the urine acquisition, the urine already collected was considered “pre-symptomatic,” and a repeat urine sampling was scheduled at least 2 weeks later following the same method.

### Sample Collection

We collected urine samples once the following criteria were met 1) no USQNB-IC symptoms for 72 hours prior to urine collection; 2) no active menstruation; and 3) no sexual activity for 72 hours prior to urine collection. A 50-100 ml urine sample from a new, unused intermittent catheter was obtained under sterile conditions. After collection, urine samples were stored at 4°C until processing. The urine was then aliquoted for use in multiple assays. One aliquot was sent to the local clinical microbiology laboratory within 8 hours of collection, during which time it was kept at 4 °C. Urinalysis (UA), urine microscopy, and standard urine culture (SUC) were completed used standard laboratory techniques.

A second aliquot was sent within 6 hours to a local laboratory for preparation for 16S rRNA gene sequencing. At the lab, urine samples were centrifuged at 4°C, 5,000 x g for 20 minutes. The supernatant was aliquoted in 2 ml cryotubes and frozen at -20°C. 10 ml PBS was added to the pellet with the remaining supernatant and then centrifuged (5,000 x g) at 4°C for 20 minutes. The pellets and aspirated PBS wash solution were stored at -20°C. Pellets were later transferred to a partner lab for DNA isolation and 16S rRNA gene sequencing.

#### DNA Isolation

Depending on the size of the pelleted material, genomic DNA was isolated either with the DNeasy Kit (Qiagen) using manufacturer’s protocol for Gram-negative bacteria or with the QIAmp DNA Micro Kit (Qiagen) using manufacturer’s protocol for DNA isolation from urine. Purified DNA was quantified using NanoDrop spectrophotometer (Thermo Fisher Scientific). Fractions of human and bacterial DNA in each sample was determined using Femto Human and Femto Bacterial DNA quantification kits (Zymo Research) according to manufacturer’s instructions.

### Sample Processing

#### Urinalysis (UA)

Urine samples were assessed by standard clinical microbiology semiquantitative chemical testing using commercial disposable test strips for glucose, bilirubin, ketone, specific gravity, blood pH, protein, urobilinogen, nitrite, and leukocyte esterase.

#### Microscopy

After centrifuging for 5 minutes, microscopic examination for and quantification of white blood cells, red blood cells, epithelial cells, yeast, bacteria, *Trichomonas vaginalis*, sperm cells, mucous filaments and crystals was performed using standard microbial techniques. UA summaries included nitrites (positive/negative), level of leukocyte esterase, and white blood cell counts (UA results are described fully elsewhere(26)).

#### Standard Urine Culture (SUC)

For SUC, a calibrated loop designed to deliver a known volume (1 mL) of urine was used. First, the sample was mixed thoroughly and the top of the container removed. The calibrated wire-inoculating loop was flamed and allowed to cool. The loop was inserted into the urine sample vertically and the loopful of urine was inoculated onto a MacConkey agar plate. A second loopful was collected and inoculated onto a blood agar plate. The plates were incubated aerobically at 35-37°C for at least 24 hours. The colonies of suspected pathogens were counted, and CFU/mL determined. Species identification was performed using Matrix-Assisted Laser Desorption/Ionization – Time-Of-Flight Mass Spectrometry (MALDI-TOF MS).

#### Genome extraction

Depending on the size of the pelleted material, genomic DNA was isolated either with the DNeasy Kit (Qiagen) using manufacturer’s protocol for Gram-negative bacteria or with the QIAmp DNA Micro Kit (Qiagen) using the manufacturer’s protocol for DNA isolation from urine. Purified DNA was quantified using NanoDrop spectrophotometer (Thermo Fisher Scientific). Fractions of human and bacterial DNA in each sample were determined using Femto Human and Femto Bacterial DNA quantification kits (Zymo Research), according to manufacturer’s instructions.

#### 16S rRNA gene sequencing

The V4 region of the 16S rRNA gene was amplified using primers 5’-TCGTCGGCAGCGTCAGATGTGTATAAGAGACAGGTGCCAGCMGCCGCGGTAA-3’ and 5’-GTCTCGTGGGCTCGGAGATGTGTATAAGAGACAGGACTACHVGGGTWTCTAAT-3’ and the following reagent concentrations: 20 mM Tris-HCl (pH 8.4), 50 mM KCl, 1.5 mM MgCl_2_, 200 µM of each dNTP, 2 µM of each primer, 1% glycerol, 0.3 U AccuPrime Taq polymerase (Thermo Fisher Scientific) and 25 ng of template DNA in 20 µl total volume. Amplification conditions were 2 minutes at 95°C initial denaturation followed by 30 cycles of 20 seconds denaturation at 95°C, 15 seconds annealing at 55°C and 5 min. extension at 72°C, and a 5-minute final extension at 72°C. Amplification products were purified with the AMPure XP system (Beckman Coulter), and their size was verified with the DNA 1000 Kit (Agilent). Indexing and pooling of amplification products was carried out according to Illumina’s 16S Metagenomic Sequencing Library Preparation protocol. The resulting library was sequenced using Illumina MySeq Reagent Kit v2 (500 cycles).

### Analysis

#### Taxonomic identification

High-quality reads were obtained by trimming low-quality sequences using Cutadapt(27). The DADA2 software(28) was then used to process the reads through filtering, dereplicating, removing chimeras, and constructing an amplicon sequence variant (ASV) abundance table. ASVs were annotated using the Bayesian LCA (BLCA) method(29) and the NCBI database. ASVs with BLCA confidence scores below 70% were labeled “unknown.” Additionally, ASVs with total counts below the sample number threshold were excluded. Contaminant ASVs were identified using control samples and were determined via a scoring method that encompasses results from Decontam(30), nonparametric statistical testing (Kruskal-Wallis p<0.5), mean comparison (i.e., mean counts of samples were less than mean counts of extraction controls), and whether sample read counts were less than the 5x greater than extraction control counts. Decontaminated ASV counts were used for downstream analysis.

#### Bioinformatic Analysis

16S results were summarized quantitatively and qualitatively (see next section). Within individuals for each time point, results were summarized with a set of alpha (within-samples) diversity indices: Observed, Evenness, Chao1, ACE, Shannon and Simpson. Observed is literally the number of ASVs uniquely identified. This index does not address the abundance of any ASV. Chao1 is an index of how many ASVs were uniquely identified, weighted by abundance to ensure that taxa present at lower abundance levels are adequately represented. ACE (Abundance-based Coverage Estimator) also reflects an abundance-based estimate of richness, but ACE includes terms to separate bacteria with greater and lesser levels of abundance in its equation, as well as a ’coverage estimator’ to more explicitly incorporate the relative abundances of even the least abundant members of the ecosystem. Larger values of these three indices mean a ’richer’ ecosystem. Evenness values, based on the Pielou equation, describe the uniformity of ASV distribution; values range from 0 to 1 and larger values indicate a more even distribution. The Shannon and Simpson indices measure both richness and evenness together; values for both range from 0-1. Shannon weights the richness by ASV evenness and larger values indicate greater diversity. Simpson emphasizes ASV evenness to a greater extent with larger Simpson values indicating less diversity(31,32), the opposite of the Shannon index. These calculations were performed in R (R 4.3.1).

Across-sample (within-individual) diversity was summarized with the Bray-Curtis dissimilarity index. Ranging from 0 (the pair of samples share all ASV) to 1 (the pair of samples share no ASVs), lower within-individual values suggest compositional consistency over time. Bray-Curtis values were computed in R (R 4.3.1) for each individual’s samples at times 1, 2, and if available, 3. We did not compute them for the 4th sample since only eight individuals had 16S rRNA gene sequencing results for this sample.

#### Urine Sample Characterization

To define normal “variability” within person, we characterized urine samples in terms of their symptoms over 72 hours, inflammatory markers, and cultivable bacteria (*i.e.*, based on Table 1, Tractenberg et al. in review 2026(26)). **Table 1** (reprinted with permission) shows the five groups we used, which are based on combinations of symptoms, UA, and SUC results. Four of the five groups describe samples where the individual remained asymptomatic for 72 hours after the sample; the fifth group is the only situation where the participant developed at least one symptom within 72 hours of the sample.

**Table 1.**
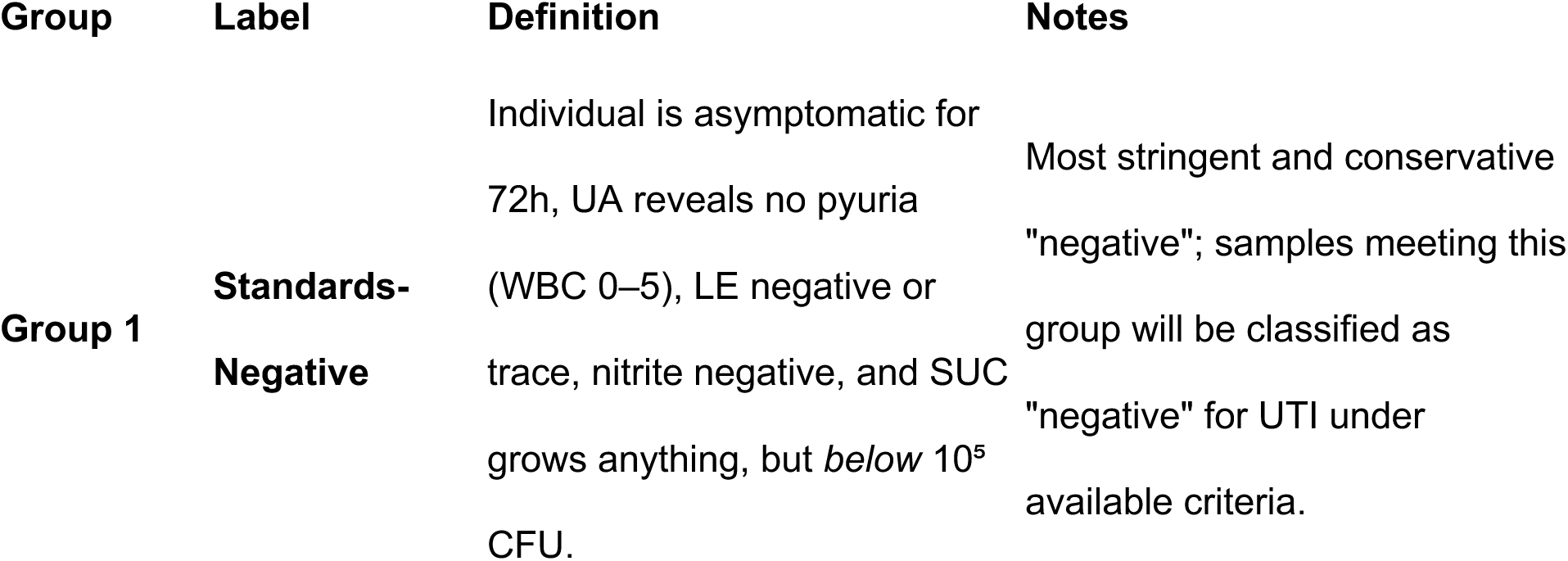
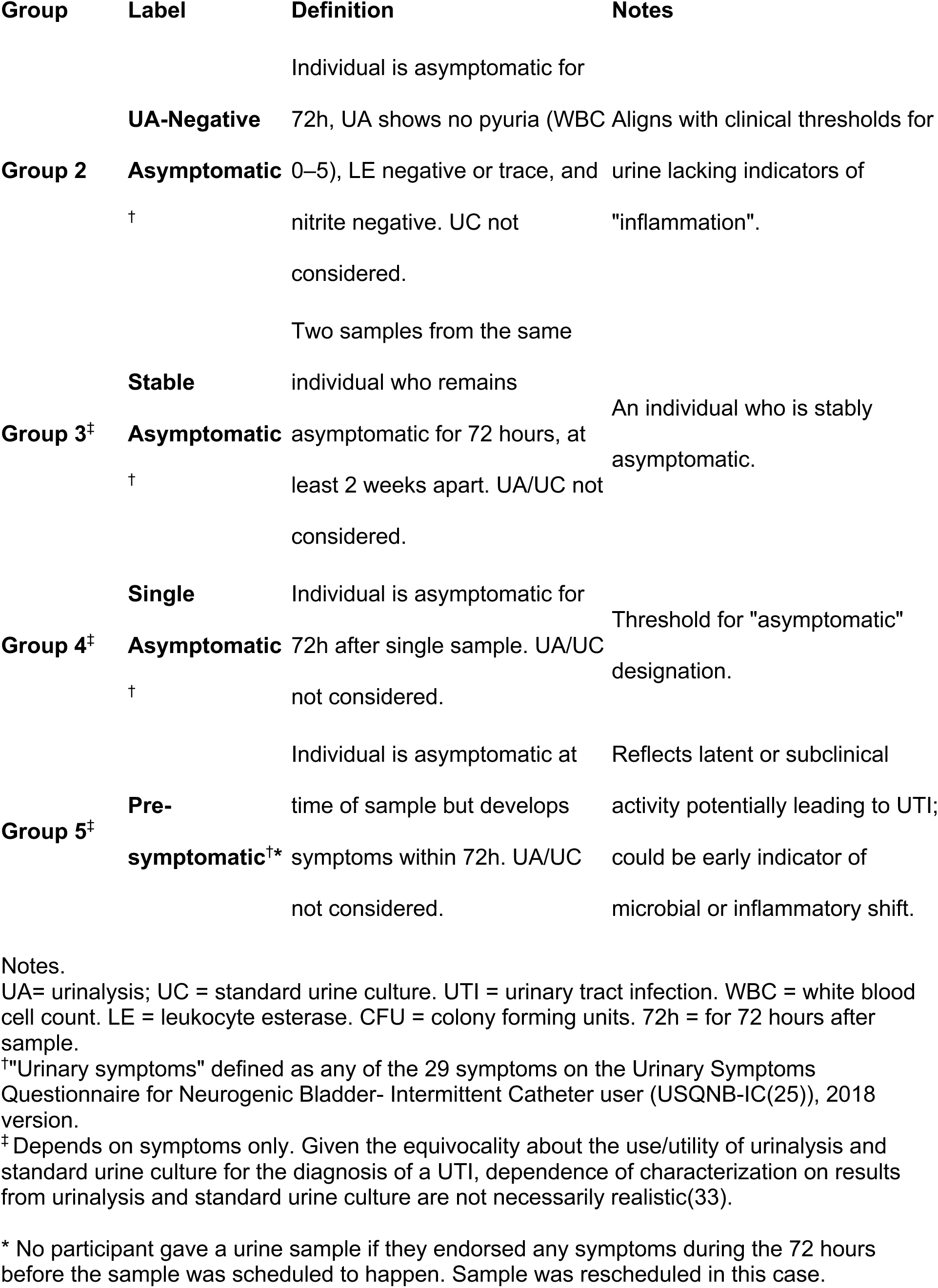
Classification of Urine Samples Based on (lack of) Symptoms, Urinalysis, and Culture in Individuals with NLUTD due to SCI/D (reprinted from Tractenberg(26) et al., in review with permission).

Only group 5 (pre-symptomatic) was mutually exclusive with all of the other groups. Each single-asymptomatic (group 4) sample could be one of consecutive asymptomatic samples; but, even if it was not part of consecutive asymptomatic samples (group 3), single-asymptomatic samples could be UA negative (group 2). Only samples that are UA negative can be standards-negative (group 1).

Data collection was performed in REDCap(34). Data analyses focused on descriptive statistics only, utilizing SPSS v. 29.0 (2024, IBM).16S summaries were calculated using R (R 4.3.1).

## Results

Each participant contributed between one and four urine samples collected at least two weeks apart while asymptomatic. Analyses focused on qualitative summaries of urotypes and quantitative summaries of alpha diversity indices to evaluate within- and between-person variability in the asymptomatic state.

The majority of individuals’ data appear at timepoints 01 and 02 because most of our participants had asymptomatic first and second samples. Some individuals developed symptoms within 72 hours of their first or second sample and so needed to provide a third and/or fourth sample. Ten individuals gave three asymptomatic samples (the third required because of laboratory errors on the 2nd one) and five individuals gave three asymptomatic samples of their four because of laboratory errors on their 3rd sample. Of the 94 participants whose samples were analyzed, 62 were male and 32 were female. All participants had SCI/D (traumatic SCI, multiple sclerosis, and spina bifida). The participants whose data are summarized in **Table 2** have been fully described elsewhere (Tractenberg et al. in review(26)).

**Table 2.** Demographic information for all participants.

| Demographics | Males<br>Mean (SD) | Females<br>Mean (SD) |
| --- | --- | --- |
| n | 63 | 31 |
| age | 45.63 (15.14) range 18-75 | 50.03 (14.60) range 23-84 |
| Yrs since dx | 15.32 (14.37) range 1-62 | 21.87 (14.03) range 1-53 |
| etiology | SCI: n = 61 MS: n=1 SB: n=1 | SCI: n = 22 MS: n=6 SB: n=3 |
| Level of Injury§ | Cervical: n= 21<br>Thoracic: n=35<br>Lumbar n=6<br>Unknown n=1 | Cervical: n= 5<br>Thoracic: n=16<br>Lumbar n=3<br>Unknown n=7 |
| Notes. dx, diagnosis; SCI, spinal cord injury; MS, multiple sclerosis; SB, spina bifida.<br>§ where injury spans levels, highest level is reported |  |  |

**Figure 1** shows that the majority of samples met the 72 hour pre- and post-sample asymptomatic criteria outlined in Table 1 and proportionally more samples from females than from males met the UA-based criteria (Group 2); otherwise, proportions of samples were generally similar in each category.

**Figure 1.**
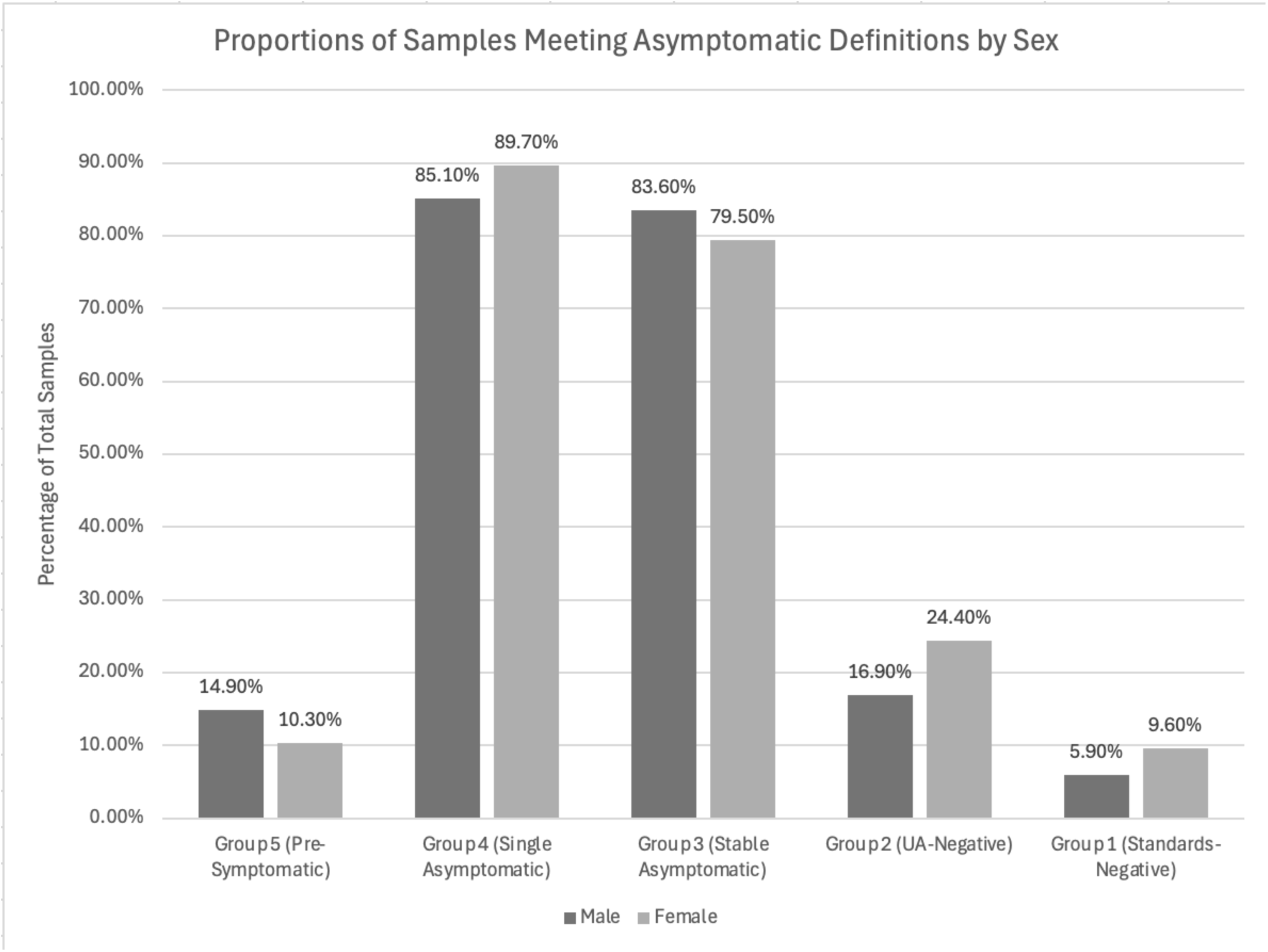
Distribution of urine samples meeting characteristics based on Symptoms, Urinalysis, and Culture. Reprinted (26)from Tractenberg et al. with permission.

**Figure 2** presents 16S rRNA gene sequencing results in the form of a series of stacked bar charts, one set for each participant. Samples are labeled according to which of the criteria outlined in **Table 1** are met. This figure presents the distribution of urotypes, defined by the predominant genus (*i.e.,* ≥50 % of total reads(8,18)), across both participants and time points. *Lactobacillus*, *Corynebacterium,* and *Gardnerella* urotypes were not detected. Instead, the most prevalent urotypes were predominated by genera considered to possess uropathogenic potential, even when participants remained asymptomatic for at least 72 hours before and after sampling. The most common of these urotypes was the genus *Escherichia/Shigella* (yellow), followed by the genera *Klebsiella* (peach)*, Enterococcus* (light gray), *Staphylococcus* (light blue), and *Streptococcus* (green). “No-predominant” urotypes were rare and, within the same individual, often followed or were preceded by one of the urotypes predominated by a genus with pathogenic potential.

**Figure 2.**
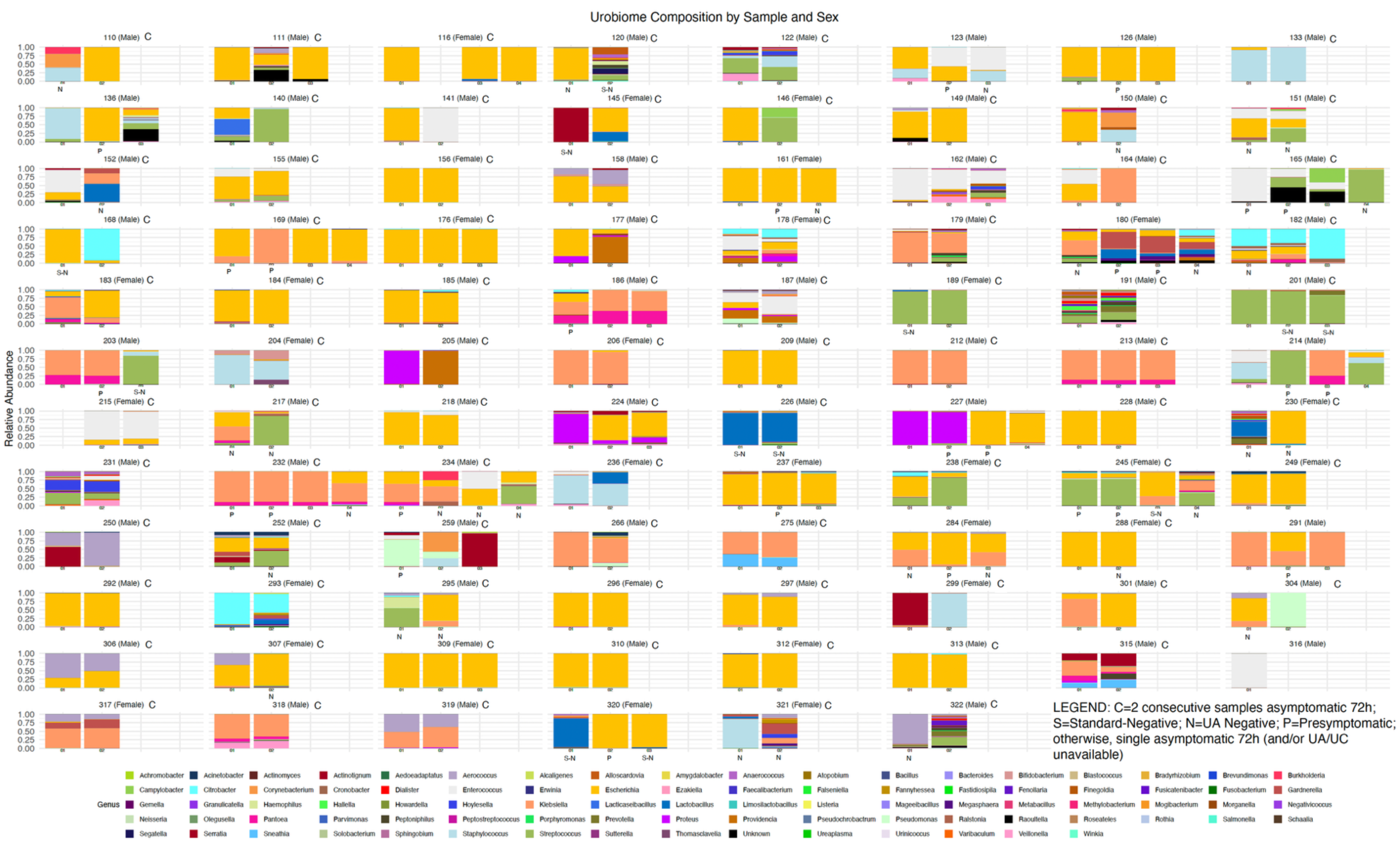
Stacked box plots showing the 87 most relatively abundant genera detected across the 229 samples contributed by the 94 participants. The category “Unknown” represents all other sequences. At the top of each set of histograms is the participant number and sex. Some are labeled **C** if at least two consecutive samples were obtained when the participant had been asymptomatic for 72 hours afterwards. At the bottom of each set of histograms, each sample is labeled using the following scheme: **P** (presymptomatic) if the individual was symptomatic within 72 hours afterwards; **N** (negative, UA) if the sample was UA-negative; and **S-N** (SUC and UA negative) if the sample was both SUC-negative and UA-negative. Samples without a designation were asymptomatic for 72 hours (single occurrence), and UA and/or SUC results may not have been available for **S or N** identification. Some individuals had to provide an extra sample (due to laboratory error) but were asymptomatic for 72 hours on three (rather than our required two) samples. One participant gave only one sample before the study ended.

Quantitative analyses of alpha (within-diversity provided further evidence that the urobiome in this population exhibits substantial normal variability (**Figure 3**). Diversity metrics included the observed number of taxa, Chao1, ACE, Evenness, Shannon, and Simpson indices. Observed, Chao1, and ACE are measures of richness, with observed being the number of detected ASVs, while both Chao1 and ACE estimate abundance-based richness with differing sensitivities to low-abundance taxa; larger values indicate greater richness. Evenness measures the distribution of ASVs; larger values indicate a more balanced community. Shannon and Simpson integrate richness and evenness, with higher Shannon and lower Simpson values denoting greater diversity.

**Figure 3.**
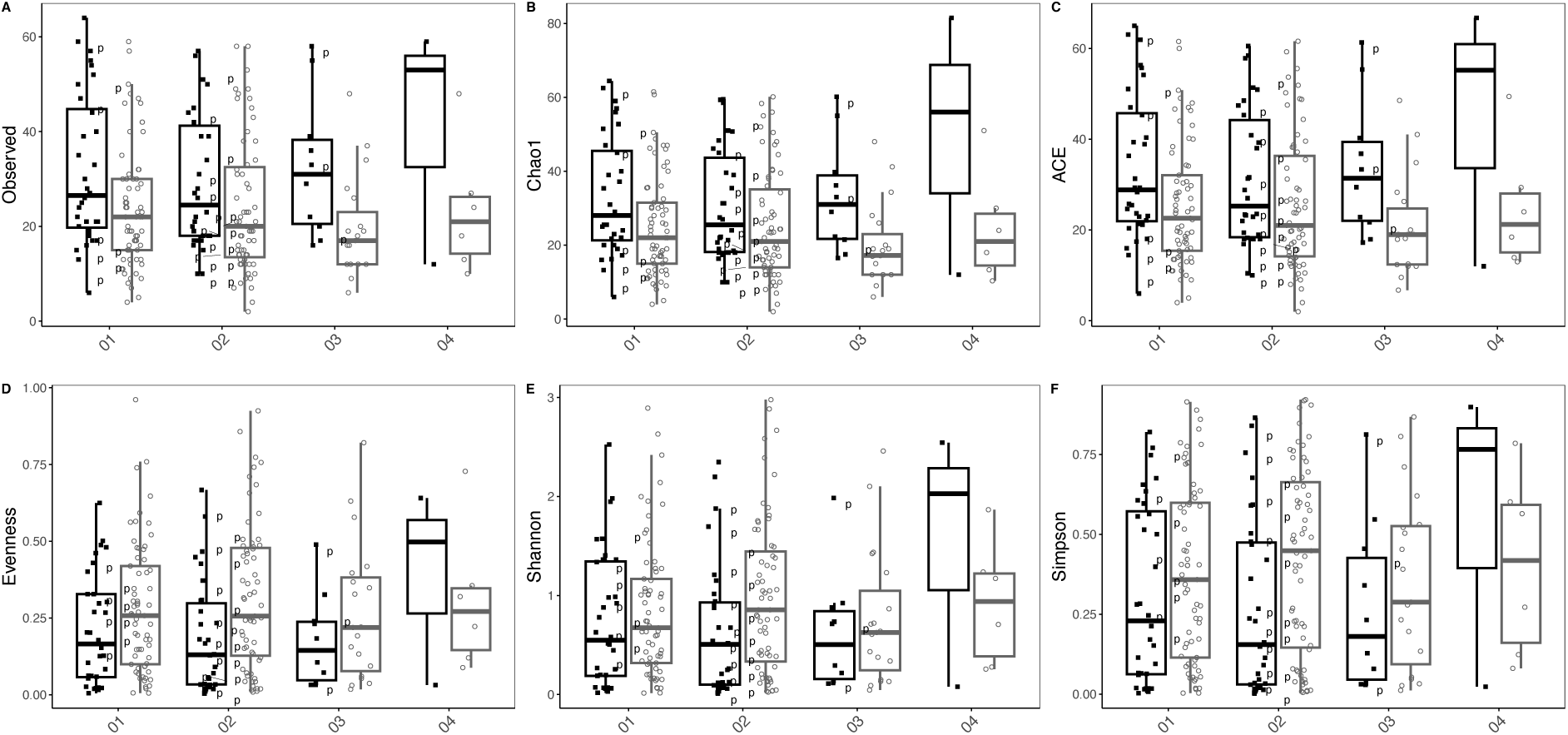
Alpha diversity indices for all samples over time. For all participants, samples are plotted in temporal order, and those collected within 72 hours of symptom onset were marked with a “p.” Samples where an individual was asymptomatic for 72 h after sample are not marked. None of the 4th set of samples met this criterion. Where a dot or a “p” appears in **black** = Samples from female participants (first plot in every panel at every timepoint), where a dot or a “p” appears in **grey**= Samples from male participants (second plot in every panel at every timepoint).

Across 94 participants, alpha diversity values spanned the full possible range for each index, regardless of sex or subsequent symptom development. Adult males exhibited slightly lower distributions of Chao1 and Shannon indices than females, although these distributions overlapped substantially. Diversity distributions differed by index and by sex among asymptomatic participants. In men, Evenness and Simpson values spanned the full range from 0 to 1, and Shannon indices ranged from 0 to 3. In women, Evenness, Simpson, and Shannon indices showed a slightly narrower—but still broad—distribution. In contrast, richness-based measures (Observed number of taxa, Chao1, and ACE) were higher and more variable in females than in males. No consistent pattern was observed between alpha diversity values and asymptomatic versus pre-symptomatic samples, with both high and low diversity values present in each group.

To assess within-person changes in microbial community composition over time, Bray–Curtis dissimilarity indices were computed for each available pair of longitudinal samples (1 vs 2, 2 vs 3, 1 vs 3) separately for males and females (**Figure 4**). Values of 1 represent completely distinct community membership, whereas 0 indicates identical taxa across the paired samples. Female participants tended to exhibit lower distances when both samples were asymptomatic and higher distances when both were presymptomatic, whereas males showed wide variability and bimodal distributions independent of symptom status. Across all comparisons, dissimilarity values ranged from 0 to 1, demonstrating substantial within-person variability in microbial composition over time.

**Figure 4.**
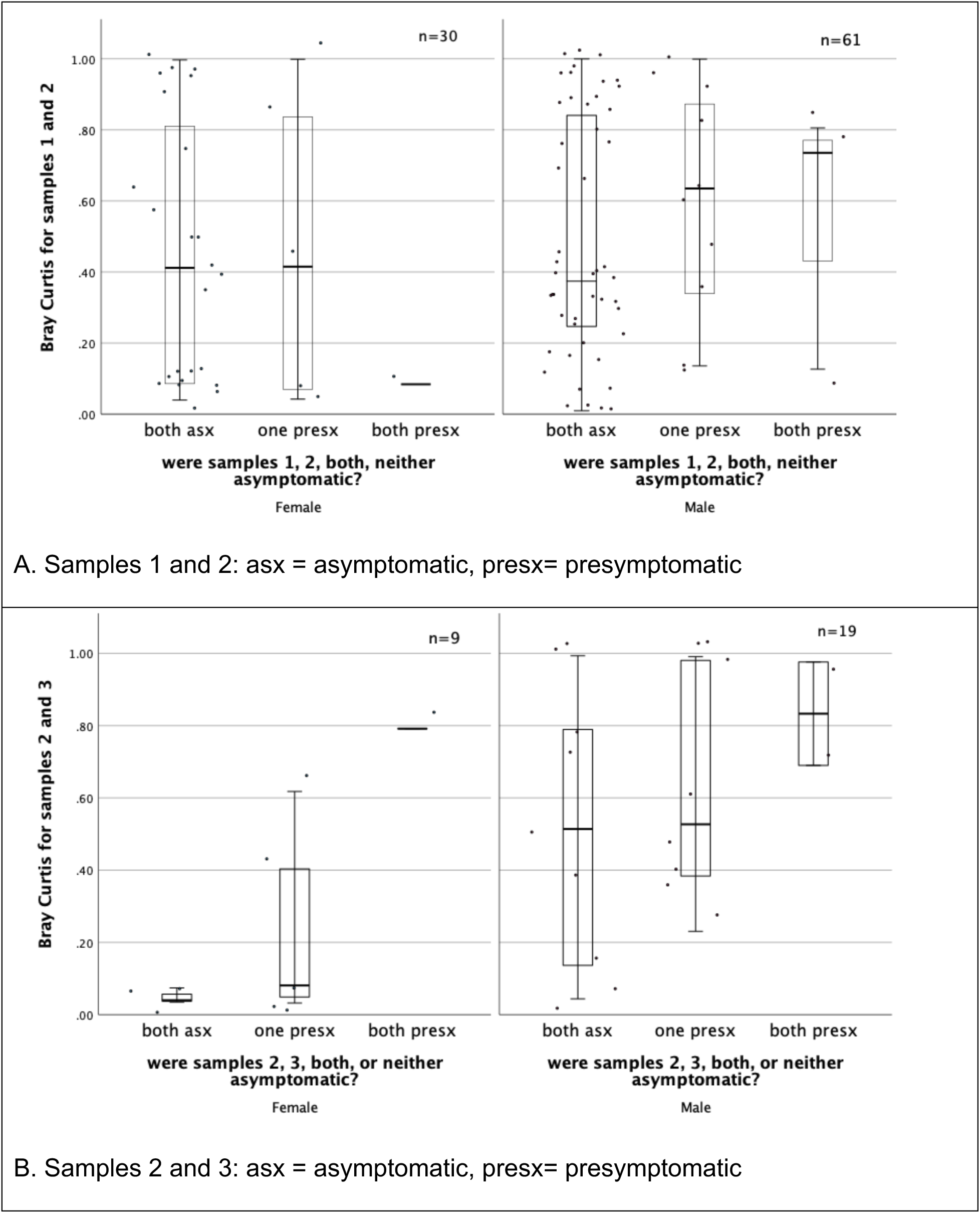
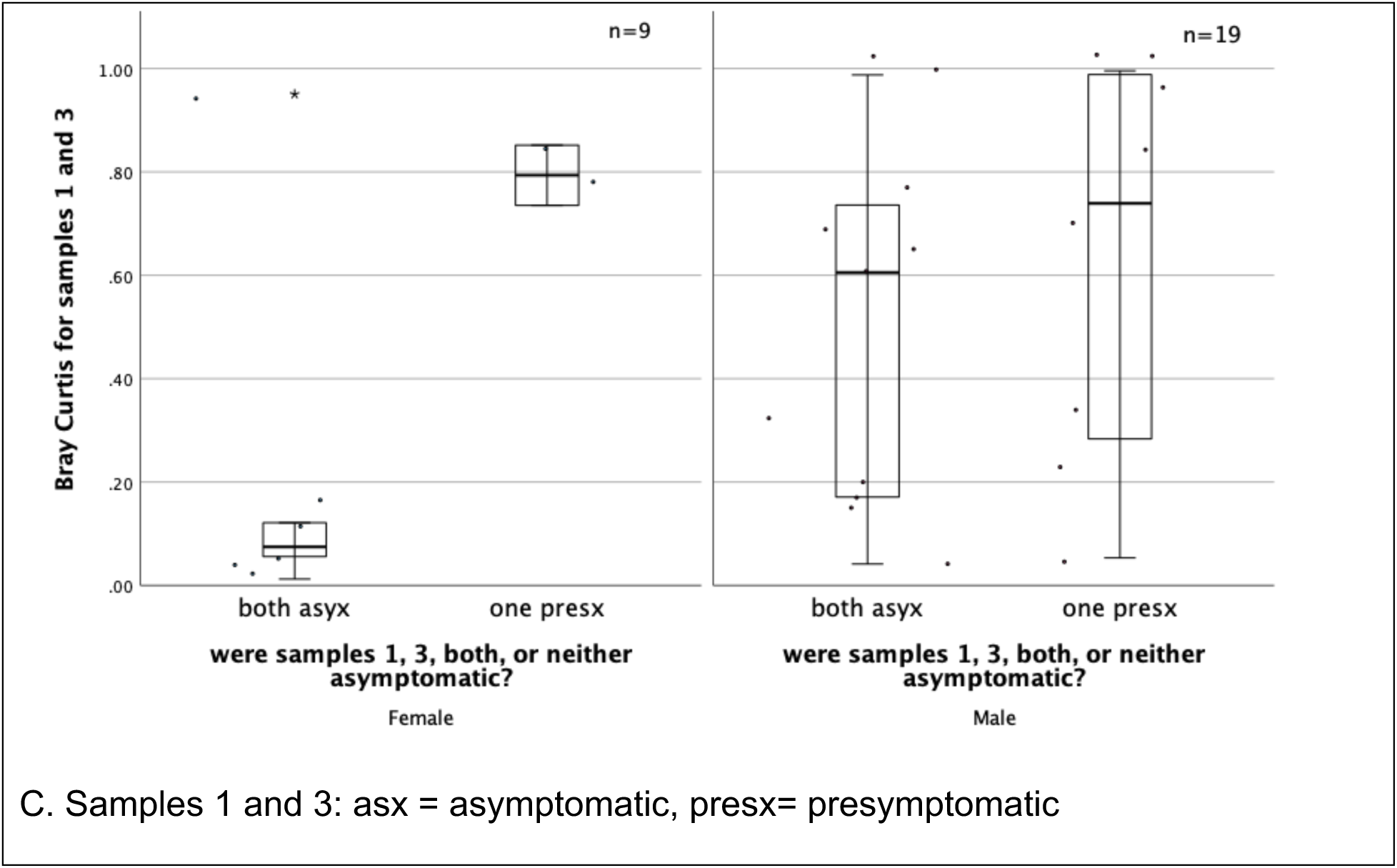
Bray–Curtis dissimilarity between paired urobiome samples from male and female participants. Panels A–C show Bray–Curtis distances between pairs of samples (A: 1 vs 2; B: 2 vs 3; C: 1 vs 3). Values near 1 indicate completely different taxa across the two samples, whereas 0 indicates identical community membership. Boxes represent the interquartile range with median lines; individual points show participant-level values. “asx” = asymptomatic for 72 hours before and after the sample; “presyx” = presymptomatic within 72 hours after the sample.

Among female participants, Bray–Curtis values tended to be lowest when both samples were collected during asymptomatic periods and highest when both were presymptomatic, with intermediate values when one of the paired samples preceded symptoms. In males, Bray–Curtis values for the first two samples were distributed bimodally—some near 0.2 and others above 0.6—regardless of symptom status, suggesting two broad patterns of temporal change rather than a monotonic relationship with symptoms. Comparisons that included a third sample (2 vs 3 or 1 vs 3) revealed extensive variability for both sexes, with values spanning most of the 0–1 range even when both samples were asymptomatic. These findings indicate that within-person microbial community composition fluctuates substantially over time and that the magnitude of this variation is not consistently associated with sex or symptom onset within 72 hours.

## Discussion

This study characterized the urobiome among individuals with neurogenic lower urinary tract dysfunction (NLUTD) due to spinal cord injury or disease (SCI/D) who manage their bladders with intermittent catheterization and who were asymptomatic at the time of sampling, across repeated measures. By analyzing repeated 16S rRNA gene sequence profiles collected during asymptomatic periods, we evaluated whether urobiome features that have been characterized as “healthy” or “potentially pathogenic” in prior cross-sectional studies could be replicated within individuals over time. Our results show that they could not. Assumptions derived from one-time group comparisons—such as predominance of certain genera indicates health or disease, or that diversity indices predict urinary health—were not supported. Instead, we observed wide intra- and inter-individual variability, substantial overlap between asymptomatic and pre-symptomatic states, and a consistent predominance of genera conventionally labeled as “uropathogens” even in the absence of symptoms(35). It is not clear whether the observed variability is due to the presence of NLUTD in our participants, to time between observations, or a combination of both. These findings collectively challenge long-standing conceptual, methodological, and interpretive assumptions about the urobiome in NLUTD, echoing a recent critique that urobiome research has yet to establish definitive causal or diagnostic frameworks(36).

## Predominance of potential uropathogens in asymptomatic urobiomes

Among these participants with NLUTD using intermittent catheterization, asymptomatic urobiomes were commonly predominated by genera often described as uropathogenic, including *Escherichia/Shigella*, *Klebsiella*, *Enterococcus* and *Staphylococcus.* These findings contrast with prior studies that reported “healthy” urobiomes are often predominated by *Lactobacillus* in adult females(18) and by Corynebacterium in males without NLUTD(37). In our cohort, *Lactobacillus*-predominant profiles were essentially absent among asymptomatic females, and *Corynebacterium* predominance was not observed in men. Instead urotypes predominated by taxa typically considered to be pathogenic were nearly ubiquitous even when participants remained symptom-free for ≥72 hours before and after sampling, suggesting that colonization by these taxa is a normal, stable feature of the urobiome in this population rather than an indicator of pathology. These observations align with accumulating evidence that bacterial species historically regarded as uropathogenic are common constituents of the urobiome among individuals with NLUTD due to SCI/D and may play context-dependent or even commensal roles(3,38). They also reinforce the argument that microbial presence and abundance alone are insufficient to define infection or health status(39). These patterns suggest that community structures can include genera traditionally regarded as pathogenic in asymptomatic individuals, and that apparent shifts between urotypes may occur independently of symptoms. Thus, for people with NLUTD due to SCI/D, the mere detection of potential uropathogens cannot be equated with disease and should not automatically trigger antimicrobial intervention. This is also true for post-menopausal females(40,41). This result has direct implications for diagnostic stewardship: microbial sequencing results must be interpreted in the context of individual clinical trajectories and symptom patterns rather than in comparison with population-level “healthy” norms derived from non-comparable groups.

## Temporal and inter-individual variability

Extending this observation, longitudinal analyses revealed that microbial community composition fluctuates substantially over time, even within asymptomatic states. Urotypes frequently shifted between collections, including transitions among uropathogen-predominant profiles without accompanying symptom changes. Bray–Curtis dissimilarities between sequential samples spanned nearly the full 0–1 range, underscoring that distinct community configurations can exist within the same person across short intervals. These results directly contradict the implicit assumption in cross-sectional analyses that between-group differences reflect stable within-person characteristics. Instead, our findings support an ecological view of the urobiome as a dynamic, adaptive system exhibiting both transient and persistent elements—a pattern consistent with longitudinal observations from prior work(2,14). In this framework, “health” reflects resilience and recovery to an individual’s baseline configuration rather than conformity to a single taxonomic state. These findings highlight the importance of establishing empirically derived reference ranges for normal variability before deviations can be meaningfully interpreted as abnormal.

## Sex-specific patterns in urobiome composition and diversity

Sex differences in the urobiome were subtle and metric-dependent. Asymptomatic males exhibited greater variability in community evenness and diversity indices, whereas females demonstrated higher richness and broader distributions of Chao1 and ACE values. These findings are consistent with prior reports that sex-linked anatomic and hormonal factors influence urobiome ecology(42), but they also reveal that such differences are quantitative rather than qualitative. Neither sex showed consistent associations between diversity metrics and symptom onset, and both displayed broad within-person variability over time. The implication is that male and female urobiomes in NLUTD are organized around comparable ecological principles—dynamic equilibrium and functional redundancy—rather than fixed, sex-specific community structures. Recognizing these subtle but consistent differences will be critical for developing analytic frameworks that distinguish biological from methodological variation in future studies.

## Methodological and interpretive implications

The failure of cross-sectional predictions of unhealthy states based on urotype or uropathogen predominance to hold under longitudinal examination exposes fundamental methodological weaknesses in the current urobiome literature. Group-level summaries and majority-rule classifications obscure intra-individual variability, while compositional data constraints inflate apparent differences between groups. The use of diversity indices as proxies for health assumes that higher diversity is inherently beneficial, an assumption that lacks empirical support in urobiome research. Indeed, systematic reviews have underscored that methodological heterogeneity and cross-sectional study designs limit interpretability in studies of lower urinary tract dysfunction(43). Our data demonstrate that every diversity level occurs in asymptomatic samples and that multivariate dimension-reduction techniques (e.g., Principal Component Analysis and Sparse Partial Least Squares Discriminant Analysis) can remove biologically relevant information if applied uncritically. Future work should therefore prioritize designs and analytic methods capable of capturing temporal dynamics—such as repeated measures, network modeling, or hidden Markov models—over cross-sectional contrasts of mean relative abundances.

## Clinical and stewardship relevance

These findings have direct consequences for both diagnostic and antibiotic stewardship. Diagnostic stewardship demands that clinical decisions be guided by data that accurately distinguish pathological change from normal variation. In the urobiome, our results confirm that a predominance of traditionally defined uropathogens does not necessarily signify infection(35), as UTI requires the presence of symptoms. We also found that high or low diversity values are not diagnostically informative since the full range of values were observed in individuals without symptoms. Without longitudinal baselines, there is a risk of labeling normal variability as disease. These results support guidance from the Infectious Diseases Society of America(44) to not routinely screen populations at high risk for recurrent UTI, such as those with NLUTD due to SCI/D with urine culture, as it is known that potential uropathogens may be present in the urine in the absence of symptoms. Doing so may expose people unnecessarily to antibiotics. This understanding is critical to antibiotic stewardship, of which a key element is avoidance of overtreatment of asymptomatic bacteriuria, particularly in populations such as NLUTD where colonization is common. Recent reviews have emphasized that progress in defining the urobiome’s clinical relevance depends on disentangling colonization from infection(8), which our findings support. By defining the empirical range of normal urobiome variability, this study provides the reference context necessary to interpret urobiome findings responsibly and to reduce inappropriate antimicrobial use.

## Limitations and future directions

Several limitations merit consideration. Although the study included 94 participants and multiple time points, the number contributing three or four samples was smaller, which may limit detection of longer-term trends. Sampling occurred at intervals of at least two weeks, leaving short-term fluctuations unobserved. Sequencing was limited to genus-level taxonomic identification; deeper metagenomic or functional analyses could reveal additional insights into metabolic or ecological stability. None of our participants developed a UTI (no matter what criteria are used), so our identification of “presymptomatic” states might not reflect true precursors to clinically significant symptoms. Despite these constraints, the consistency of patterns across multiple indices and subgroups strengthens the conclusion that observed variability reflects biological rather than technical noise; therefore, these repeated measures successfully characterize the range of normal variability. Future studies with higher-frequency sampling and species- or strain-level resolution will be essential to model transitions, resilience, and recovery dynamics more precisely. Next steps may also include functional assessments of these various urobiomes to better determine the functional roles of both potentially uropathogenic and commensal species.

## Conclusions

Within and across individuals, considerable variability was observed in 16S rRNA gene sequencing results in the absence of urinary symptoms. Asymptomatic states commonly feature potential uropathogens, diversity metrics fail to distinguish health from pre-symptomatic states, and intra-individual variability is substantial. These findings underscore the necessity of redefining “normal” in terms of variability and resilience rather than static composition. Establishing this empirical baseline is a prerequisite for reliable diagnostic interpretation and for advancing both diagnostic and antibiotic stewardship in neurogenic bladder management.

## Data Availability

All data produced in the present study are available upon reasonable request to the authors

